# ESTIMATING R_0_ OF SARS-COV-2 IN HEALTHCARE SETTINGS

**DOI:** 10.1101/2020.04.20.20072462

**Authors:** Laura Temime, Marie-Paule Gustin, Audrey Duval, Niccolò Buetti, Pascal Crépey, Didier Guillemot, Rodolphe ThiéBaut, Philippe Vanhems, Jean-Ralph Zahar, David R.M. Smith, Lulla Opatowski

**Author notes:** Corresponding author: Laura Temime, MESuRS laboratory, Conservatoire national des Arts et Métiers, 292 rue Saint-Martin – 75141 Paris Cedex 03, France. On behalf of the “Modelling COVID-19 in hospitals” REACTinG AVIESAN working group: Niccolò Buetti, Christian Brun-Buisson, Sylvie Burban, Simon Cauchemez, Guillaume Chelius, Anthony Cousien, Pascal Crepey, Vittoria Colizza, Christel Daniel, Aurélien Dinh, Pierre Frange, Eric Fleury, Antoine Fraboulet, Didier Guillemot, Marie-Paule Gustin, Bich-Tram Huynh, Lidia Kardas-Sloma, Elsa Kermorvant, Jean Christophe Lucet, Lulla Opatowski, Chiara Poletto, Laura Temime, Rodolphe Thiebaut, Sylvie van der Werf, Philippe Vanhems, Linda Wittkop, Jean-Ralph Zahar.

## Abstract

To date, no specific estimate of R_0_ for SARS-CoV-2 is available for healthcare settings. Using inter-individual contact data, we highlight that R_0_ estimates from the community cannot translate directly to healthcare settings, with pre-pandemic R_0_ values ranging 1.3-7.7 in three illustrative healthcare institutions. This has implications for nosocomial Covid-19 control.

---

In the context of the current Covid-19 pandemic, the basic reproduction number R_0_ has been recognized as a key parameter to characterize epidemic risk and predict spread of SARS-CoV-2, the causative virus of Covid-19 infection [1]. R_0_ describes the average number of secondary cases generated by an initial index case in an entirely susceptible population. R_0_ is determined not only by the inherent infectiousness of a pathogen, but also environmental conditions, host contact behaviours and other factors that influence transmission. Understanding the evolution of the effective reproduction number R_t_, which describes R_0_ as it varies over time, is also essential for epidemiological forecasting and to assess the impact of control strategies [2, 3].

Over recent months, numerous estimates of R_0_ for SARS-CoV-2 have been computed through analysis of reported infections from countries all over the world [2, 4-6], as well as in specific subpopulations, such as individuals aboard the Diamond Princess cruise ship [7]. Published estimates mostly range from 2-4.

However, to date, no estimates of R_0_ specific to healthcare settings have been published.

Healthcare institutions are confronted with several urgent and overlapping challenges linked to Covid-19. Acute care facilities face unprecedented demand for beds and resources to accommodate Covid-19 patients, particularly in intensive care units in high-prevalence regions. Introduction of SARS-CoV-2 to healthcare settings can further result in nosocomial outbreaks, with superspreading events already reported in some hospitals [8], as was also observed for SARS-CoV and MERS-CoV. In addition to risks for patients, whose underlying conditions put them at greater risk of severe infection, there is also an important risk of infection among healthcare workers [8].

Contacts between individuals are fundamental to the spread of respiratory pathogens like SARS-CoV-2, and contact patterns in healthcare settings are highly context-specific. Contacts between patients and healthcare workers tend to be simultaneously more frequent, longer and more at-risk than contacts occurring in the community. This could translate to higher R_0_ values, as underlined in earlier work on other coronaviruses, in which R_0_ was estimated to be much higher in hospitals than in the community [9].

Here, using detailed individual-level contact pattern data from both the community and three healthcare institutions in France, we explore how the reproduction number estimated in the community translates to these institutions, and discuss potential consequences for public health.

## METHODS

Under simplifying assumptions, R_0_ can be estimated as follows:

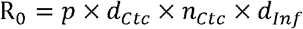

where *p* is the probability of transmission per minute spent in contact, *d*_*Ctc*_ is the average contact duration (in minutes), *n*_*Ctc*_ is the average number of contacts per person per day, and *d*_*Inf*_ is the average duration of infectivity (in days): approximately 10 days for Covid-19 [10].

Assuming that *p* and *d*_*Inf*_ are the same for individuals in the community and in healthcare settings, we can translate the previous expression into setting-specific R_0_ values computed as:

- In the community: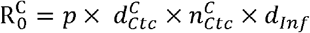
- In the healthcare settings: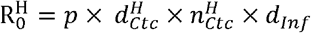

where superscripts *C* and *H* denote values for community and healthcare settings, respectively.

The healthcare setting-specific reproduction number may then be estimated from the community-specific reproduction number and the contact pattern characteristics in both settings, as:

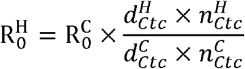

## NUMERICAL APPLICATION IN THE FRENCH CONTEXT

Based on detailed inter-individual contact data from France [11], in the community the median number of inter-individual contacts per person is 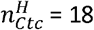 contacts/day and the median duration of these contacts ranges from 15 minutes to 1 hour. For simplicity, in the following we use 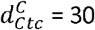 minutes.

The reproduction number for SARS-CoV-2 has been estimated in the French community at values ranging from 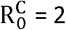 to 4 [2, 12, 13]. In the following we use 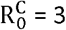.

These translate to an average transmission risk per minute spent in contact of:

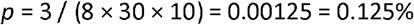

Table 1 provides estimates of the healthcare setting-specific reproduction number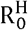, depending on the average number of daily contacts within the healthcare setting 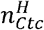, and the actual value of 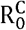. The mean duration of daily contacts within the healthcare setting 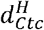 is assumed to range from 10 to 40 minutes.

**Table 1.** Range of estimated reproduction numbers 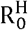 values obtained when 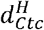 ranges from 10 to 40 minutes, for different assumed values of 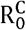 (rows) and 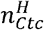 (columns)

| | | Average number of daily contacts in the healthcare setting ($n_{ctc}^H$) | | | | |
| --- | --- | --- | --- | --- | --- | --- |
|  |  | 5 | 10 | 15 | 18 | 20 |
| Assumed value for<br>basic reproduction<br>number in the<br>community ($R_0^C$) | 2 | 0.4-1.7 | 0.8-3.3 | 1.3-5 | 1.5-6 | 1.7-6.7 |
|  | 2.5 | 0.5-2.1 | 1-4.2 | 1.6-6.3 | 1.9-7.5 | 2.1-8.3 |
|  | 3 | 0.6-2.5 | 1.3-5 | 1.9-7.5 | 2.3-9 | 2.5-10 |
|  | 3.5 | 0.7-2.9 | 1.5-5.8 | 2.2-8.8 | 2.6-10.5 | 2.9-11.7 |
|  | 4 | 0.8-3.3 | 1.7-6.7 | 2.5-10 | 3-12 | 3.3-13.3 |

### THREE ILLUSTRATIVE EXAMPLES

As an illustration, we used detailed contact data from three different healthcare settings in France during the pre-pandemic period to estimate 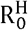 in the absence of control measures specific to Covid-19:

- For a 170-bed rehabilitation hospital [14], where 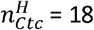 contacts/day and 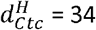 min, the pre-pandemic 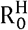 is estimated as

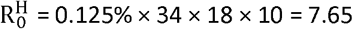
- For an acute-care geriatric unit [15], where the cumulative time spent in contact with others per individual per day was 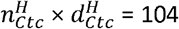, the pre-pandemic 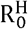 is estimated as

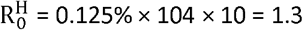
- For a 100-bed nursing home [16], where the cumulative time spent in contact per individual and per day was 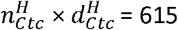, the pre-pandemic 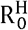 is estimated as

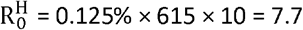

## DISCUSSION

Estimating R_0_ has been an important focus of epidemiological work to understand the transmission dynamics and pandemic trajectory of SARS-CoV-2. We highlight here that reproduction numbers estimated in the community cannot be translated directly to healthcare settings, where inter-individual contact patterns are specific to and variable between institutions.

Health care institutions are at high risk of SARS-CoV-2 importation, from admission of infected patients or from visitors or healthcare workers infected in the community. Our estimates of 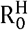 suggest that, depending on a healthcare facility’s size and structure, the risk of nosocomial spread may be much higher or lower than in the general population, with values ranging from 0.4 to 13.3 (Table 1).

Our results have implications for Covid-19 infection prevention and control. In healthcare settings with estimated low values of pre-pandemic 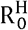, it is expected that classical barrier measures – reducing p, the probability of transmission per minute of contact – will suffice to prevent the spread of the virus. On the contrary, in healthcare settings where the estimated pre-pandemic 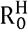 is high, it is critical to implement additional control measures. These measures could include reducing the frequency 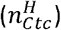 and duration 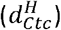 of contacts (e.g. through limiting patient-patient contacts by cancelling social activities and gatherings), limiting patient transfers, or reorganizing human resources and provisioning of care within the institution.

It should be underlined that this work’s aim is to present a conceptual discussion about R_0_ in healthcare settings. Hence, the elements presented here, and in particular the numerical estimates, should be interpreted in light of the following over-simplifications.

First, Covid-19 infection was simplified by assuming the same duration of infectivity, irrespective of the setting. However, in the community, individuals presenting symptoms may isolate themselves and stay at home whereas patients of healthcare settings will stay hospitalized. Considering such differences would lead to higher estimates of 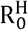.

Second, we assumed the same per-minute probability of transmission, irrespective of the setting and nature of contacts. However, some hospital contacts, such as those involving close proximity or invasive procedures, may pose greater transmission risk than others. Also, a higher concentration of severe infections, which may shed more virus [17], and the presence of immunosuppressed individuals, may entail a higher transmission probability in hospitals, therefore increasing 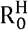.

Third, 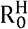 may differ according to individual characteristics, notably for patients vs. healthcare workers. In addition, some individuals may be super-contactors or super-shedders, with a greater probability of generating secondary cases if infected.

Last, our R_0_ formula assumes random homogenous mixing between individuals in the population. For hospital networks, which are highly clustered due to ward structure and occupational hierarchies, this formula could be refined. Computing 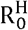 values using contact information at the ward level should facilitate more accurate estimates. Additionally, our formula makes the assumption that transmission risk increases linearly with contact duration, which may not be correct, especially for very long contacts. For instance, censoring contacts longer than 1 hour in the data from the first example gives an average contact duration within the facility of 15 min, leading to a lower estimated 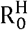 of 3.37.

In conclusion, pandemic Covid-19 continues to overwhelm healthcare institutions with critically ill and highly infectious patients, and nosocomial outbreaks pose great risk to patients and healthcare workers alike. Understanding how transmission risk varies between community and healthcare settings, and within and between different healthcare institutions such as hospitals and long-term care facilities, is fundamental to better predict risks of nosocomial outbreaks and inform appropriate infection control measures.

## Data Availability

All the data referred to in the manuscript comes from published papers.

## FUNDING

This work was funded in part by the French government through the National Research Agency projects SPHINX (# 17-CE36-0008-01) and MOD-COV. DS is also supported by a Canadian Institutes for Health Research doctoral foreign study award (Funding Reference Number 164263).

## Notes

### Competing Interest Statement

The authors have declared no competing interest.

